# An Algorithm to Predict New Magnetic Resonance Imaging Lesions to Support Improved Disease Surveillance in Multiple Sclerosis

**DOI:** 10.1101/2024.06.20.24309267

**Authors:** Michael S. Robinette, Karla Gray-Roncal, Kathryn C. Fitzgerald, Kadija Ferryman, Casey Overby Taylor, John Scott, Elias S. Sotirchos, Peter A. Calabresi, Ellen M. Mowry, William Gray-Roncal

## Abstract

**Background:** People with multiple sclerosis (MS) undergo magnetic resonance imaging (MRI) to monitor disease activity and treatment response. Current scan frequency recommendations are non-individualized, potentially increasing unnecessary imaging and costs. Risk-based tools could enable more personalized surveillance strategies.

**Objective:** To develop an algorithm that predicts new lesions in subsequent brain MRI.

**Methods:** Using longitudinal data from adults with MS at the Johns Hopkins MS Center (2017–2025) with ≥2 visits and ≥1 MRI scan, a logistic regression model with 5-fold stratified cross-validation predicted new lesions, using a sensitivity-prioritized threshold. Features included disease activity history, therapy class, and patient-reported outcomes.

**Results:** Among 1,131 participants (3:1 female-to-male; mean age 48, SD 12.3), 8.8% developed new MRI lesions. At a 0.08 threshold, sensitivity was 0.72 and specificity 0.75, with AUC 0.80. The model identified 72 patients with new lesions and 772 without (low risk). Prior MRI activity and recent relapse predicted new lesions, while older age and high-efficacy DMT use were associated with lower risk.

**Conclusion:** The algorithm accurately stratified patients by risk of MRI lesion activity and identified who could undergo longer surveillance intervals with low risk of missed inflammation. With validation and integration, it may enable personalized monitoring in MS care.

## Introduction

Multiple sclerosis (MS) is a chronic immune-mediated disease of the central nervous system in which early pathology is dominated by focal inflammatory demyelination, visualized as signal changes, or lesions, in brain and spinal cord on magnetic resonance imaging (MRI). Over time, many patients transition to secondary progressive MS, with gradual disability accumulation despite declining overt inflammatory activity. Early treatment with disease-modifying therapies (DMTs) reduces MRI lesion accrual and relapse frequency; however, substantial heterogeneity exists across therapies in their ability to suppress MRI activity^1,2^. Given the prognostic significance of MRI lesion burden and activity, treatment decisions increasingly rely on imaging-based markers to balance efficacy and safety in individualized therapeutic strategies.

Monitoring the treatment response to MS DMT has not yet been optimized, particularly in the modern treatment era. Current practice relies on routine clinical assessments and periodic MRI surveillance to detect subclinical disease activity. However, the appropriate frequency of imaging, especially for specific patients or across patient subgroups (e.g., older patients, those on higher efficacy DMT), has not been determined. Existing recommendations are broad and non-individualized, leading to potentially unnecessary imaging, patient burden, and cost, and highlighting the need for tailored monitoring strategies that incorporate patient-specific risk and disease stability^3,4^.

Statistical and machine-learning approaches allow for a systematic approach to explore various classification methods to optimize performance while emphasizing relevance to clinical practice (e.g., interpretability). These algorithms can utilize large amounts of patient data (e.g., prior imaging results, relapse history, medication information) to use for training of their prediction models, with outputs that could potentially be read by providers and used as decision-making aids for clinically relevant questions.

Our objective was to develop an algorithm to predict the presence of new lesions in a subsequent MRI scan.

## Methods

All methods were carried out in accordance with relevant guidelines and regulations. Informed consent was obtained from all subjects and/or their legal guardian(s). Our study was approved by the Johns Hopkins Medicine Institutional Review Board.

### Participants

We obtained data from participants that attended clinic visits at the Johns Hopkins Multiple Sclerosis Center between 2017 and 2025 and included those that had at least two clinic visits. We excluded those who had switched to a more aggressive DMT since their last visit, as a follow-up MRI would be indicated for these participants to establish a new baseline when changing MS therapies.

### Data Sources

Our algorithm development included data from two standardized sources: the MS Performance Test (MSPT)^5^ and the MS Smartform^6^. The MSPT is an iPad-based standardized assessment that is collected during routine visits and includes objective performance measures (walking speed, dexterity score, processing speed) as well as self-reported outcomes such as anxiety, depression, and fatigue (as subscales of NeuroQoL ^7^). The MS Smartform is a tool for MS providers to record clinical data such as relapses, number of new lesions on MRI (collected at intervals of 6-12 months), and DMT, which are readily available in the electronic health record (EHR).

For preprocessing, the MSPT assessment date served as the index clinical visit date used to align data across sources. Of the available MSPT measures, the feature set was reduced to anxiety, depression, fatigue, and the Patient-Determined Disease Steps (PDDS) score; performance-based assessments specific to the MS PATHS consortium infrastructure (e.g., dexterity and memory tests) were excluded to maximize generalizability to settings outside of PATHS-affiliated sites. From the MS Smartform, medication history was categorized into traditional or aggressive DMT classes, and only brain MRI scans were included given the distinct clinical presentation of brain versus spinal cord lesions; when both radiology and physician assessments were available, physician determination was prioritized. MRI records were linked to a given visit if they fell within a three-month window of the MSPT assessment date. Medication records were aligned by confirming that the medication start-to-stop window included the visit date.

For patients with multiple clinic visits on record, the most recent visit with a qualifying linked MRI scan was selected as the index record, ensuring each patient contributed a single independent observation.

### Classification Approach

The output of our classifier predicts if a new lesion will occur during the next scan. For classification, we used logistic regression to predict the target variable (i.e., new lesion present). Logistic regression was chosen for its ease of use, accuracy, and interpretability across a compact feature set; these qualities are important to build clinical trust and effectively deploy tools. We used a 5-fold stratified cross-validation technique, which trains and tests the data by iteratively splitting the set into 80%/20% training/test groups, resulting in each patient encounter being evaluated in the test set exactly once.

Predictor variables included patient-reported outcomes from the MSPT (anxiety, depression, and fatigue as subscales of NeuroQoL, and the PDDS score), DMT class (categorized as traditional or aggressive), relapse history (time since last relapse and number of relapses in the prior two years), presence of new lesions on the prior MRI scan, clinical visit date, age, and sex. All continuous variables were z-standardized across the cohort so that larger values corresponded to increased magnitude of each variable, including anxiety, depression, and fatigue.

Outputs from the algorithm, which we named Longitudinal, Equitable Systematic Imaging Operations for Neuroimmunology (LESION), included an outcome coefficient related to estimated probability of new lesion(s), as well as the features (variables) that contributed to that result with their corresponding strengths. In order to determine which coefficient values would be labeled as “new lesion predicted” versus “no new lesion predicted,” various thresholds were evaluated for their respective sensitivity and specificity. The operating threshold was then selected to optimize sensitivity, correctly predicting new lesions on follow-up scan (as this can pose significant clinical risk), while still maintaining relatively high specificity to limit unnecessary scans. This threshold was empirically chosen to balance these concerns, with the support of MS clinicians in our team.

## Results

### Participant Characteristics

We included data from 1131 participants for development and testing of the classifier in LESION. Their average age was 48 years with a 3:1 female-to-male ratio; 900 (80%) self-identified as White; 219 (19%) as Black; and 11 (1%) as Asian. Within this cohort, average time since last relapse was 9.1 (SD 6.7) years, 520 were on a higher-efficacy (aggressive) DMT, and 467 were on a moderate-efficacy (traditional) DMT. In aggregate, 100 (8.8%) had a new lesion on their follow-up MRI, which served as the model outcome (Table 1).

**Table 1:** Baseline characteristics of patients included in LESION development and testing (N = 1131). The cohort reflects the demographic composition of a large academic MS center, with representation across race, sex, and disease-modifying therapy class. ^+^ Alemtuzumab, Natalizumab, Ofatumumab, Ocrelizumab, Rituximab, Cladribine. ^++^ Glatiramate acetate, Interferons, Teriflunomide, Fumarates, Sphingo-1 phosphate modulators.

| **Characteristic** |  |
| --- | --- |
| Total Patients, N | 1131 |
| Age yrs, mean (SD) | 48.3 (12.3) |
| Race, N (%) |  |
| White | 900 (79.6) |
| Black | 219 (19.3) |
| Asian | 11 (0.97) |
| Female:Male ratio | 3:1 |
| DMT, N (%) |  |
| Aggressive (Higher Efficacy)<sup>+</sup> | 520 (45.9) |
| Traditional (Moderate Efficacy)<sup>++</sup> | 467 (41.3) |
| Patients w/ New Lesions, N (%) | 100 (8.8) |
| Time since relapse yrs, mean (SD) | 9.1 (6.7) |

### Algorithm Results

In order to select the operating threshold for LESION, various outcome coefficient cut-off values were tested to assess their sensitivities and specificities, which are plotted as a curve in Figure 1. Overall, the area under the curve (AUC) for our model was 0.8. The classification probability threshold of 0.08 was chosen in collaboration with MS neurologists, and had a sensitivity of 0.72 and specificity of 0.75. At this algorithm setting, we obtained 72 true positives (new lesions correctly predicted for the next scan) and 772 true negatives (no new lesions correctly predicted for the next scan), suggesting that a majority of patients (68%) in our cohort may represent candidates for extended surveillance intervals (Table 2).

**Table 2:** Confusion matrix for the LESION algorithm at the selected operating threshold. The algorithm correctly identified 72 of 100 patients with new lesions (sensitivity = 0.72) while correctly classifying 772 patients (68% of the total cohort) who did not have new lesion activity. Among patients predicted not to have a new lesion, 28 (2.5% of total cohort) were found to actually have new disease activity on subsequent MRI.

|  | Patients with New Lesions<br>N (%) | Patients without New Lesions<br>N (%) |
| --- | --- | --- |
| Predicted with New Lesion | 72 (72) | 259 (25) |
| Predicted without New Lesion | 28 (28) | 772 (75) |

**Figure 1.**
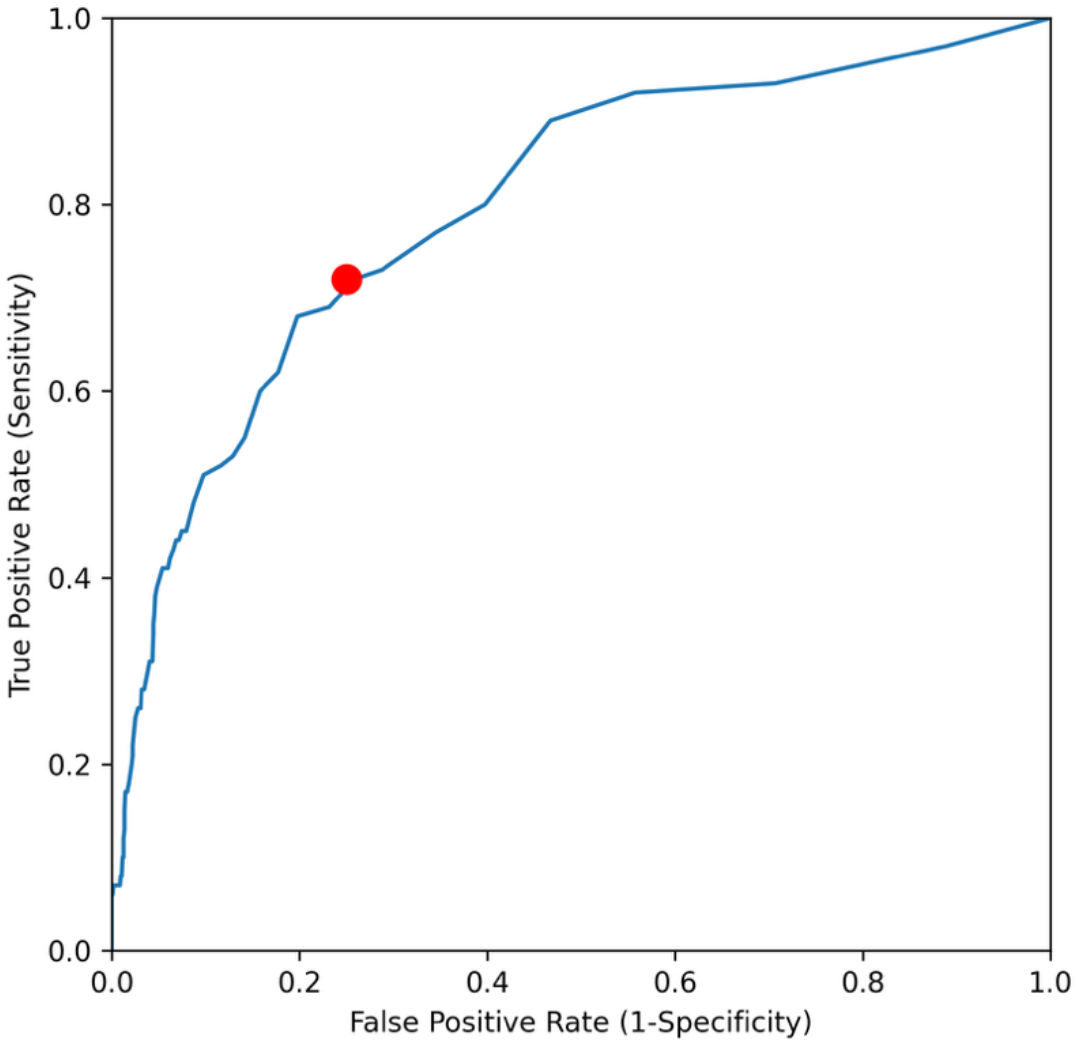
Receiver operating characteristic (ROC) curve for the LESION algorithm. Each point on the curve represents a candidate classification threshold, plotted by true positive rate (sensitivity) against false positive rate (1 − specificity). The selected operating point (red) was clinically chosen to maximize sensitivity (minimizing missed lesions) while maintaining acceptable specificity. AUC = 0.8.

As shown in Figure 2, features that were significantly associated with stronger prediction of new lesions included prior lesion at last MRI (coefficient ≈ +2) and number of relapses in the past two years, while features associated with reduced likelihood of new lesions included aggressive medication use (coefficient ≈ -2) and older age. The remaining features (visit date, sex, fatigue, anxiety, depression, PDDS) were not statistically significant.

**Figure 2.**
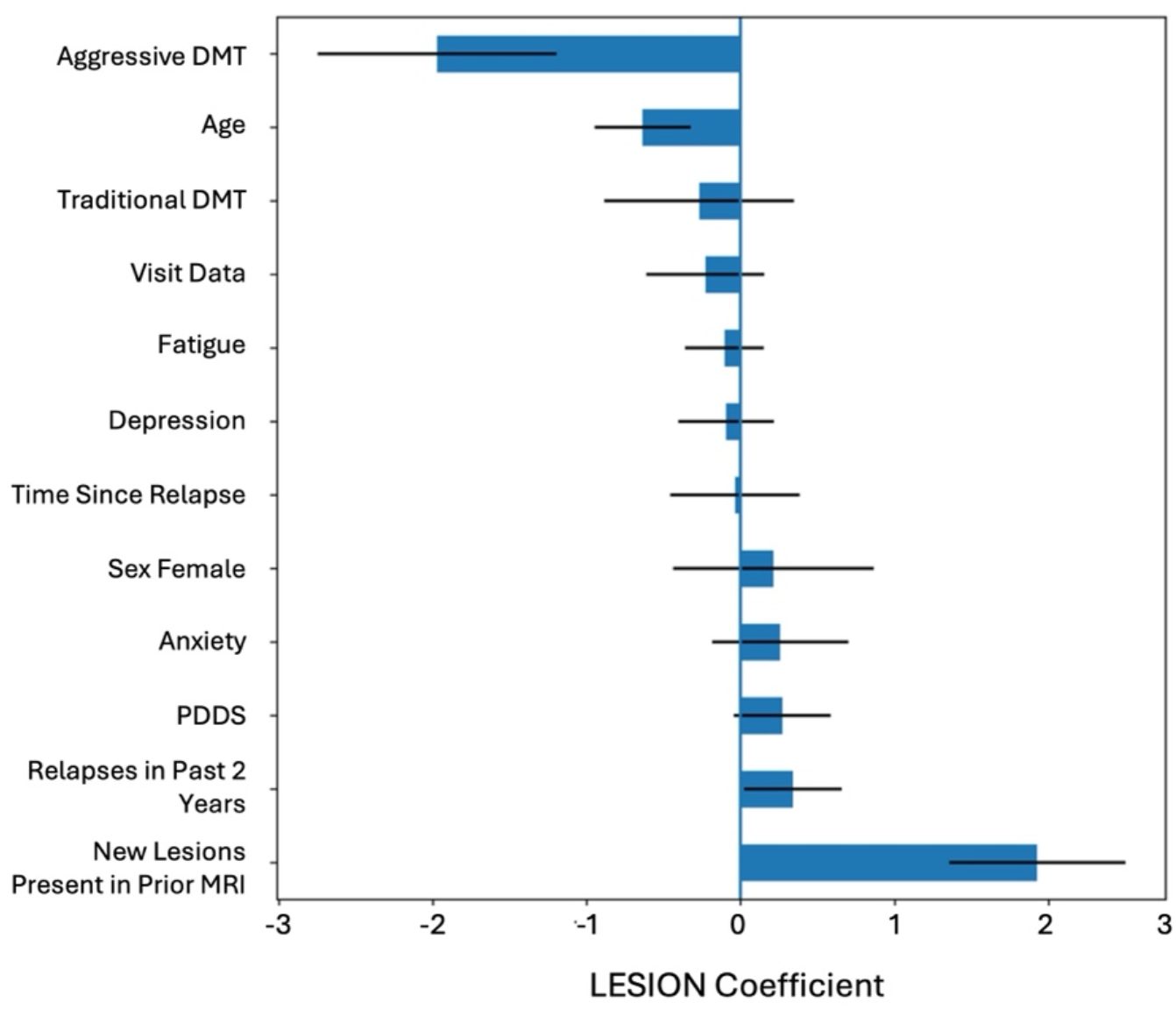
Algorithm variables or features. The relative importance of each feature can be assessed through the logistic regression coefficients. Because all continuous variables are z-standardized across the cohort, coefficients are directly comparable in magnitude (larger absolute values indicate stronger drivers of prediction). Negative coefficients indicate features associated with reduced likelihood of new lesions, while positive coefficients indicate features associated with increased likelihood. Features whose 95% confidence intervals do not cross zero are considered statistically significant contributors to the model.

## Discussion

In this initial prototype, LESION correctly identified patients with higher predicted risk of demonstrating new lesions on the next MRI. Just as important, it likewise identified many patients (68% of cohort) who could have skipped their next MRI with a low risk of missing a new lesion. If the neurologist has already been considering extending the interval between MRI scans, having the result of the LESION algorithm in hand may provide additional reassurance or a “second opinion” to support the clinician’s assessment, which is based on data specific to the patient.

The variables or features that most strongly predicted a new lesion on future MRI were congruent with findings from previous studies: older participants, those using higher-efficacy DMTs, and individuals with stable disease (no recent relapses or new lesions) were less likely to have evidence of new inflammation in the next scan. Other features included did not reach statistical significance and may warrant further study, especially with increased sample sizes and broader populations.

The potential impact of reducing unnecessary imaging extends across multiple dimensions. While not every recommendation to skip the MRI will be followed, health system costs may be reduced (e.g., a typical brain MRI might cost as much as $3500 per scan) by incorporating the LESION tool. In addition to the cost savings, the tool can target resources for those who do need more frequent monitoring. Access to MRI scans can be limited, especially in certain geographic areas, both in the United States and in other countries; furthermore, even when MRIs are available, patients often must wait months to obtain an appointment. It is also important to emphasize the benefit to the patients who can forego frequent scans, particularly to those with claustrophobia, limited mobility, or chronic back pain, for whom an MRI scan can be a difficult ordeal, as well as for those for whom copay burden posits substantial resource strain.

More complex models that consider these predictions could be used in addition to or instead of simple logistic regression, potentially at the risk of more limited interpretability (i.e., impacting clinician and patient trust) or the requirement for additional data examples (e.g., deep learning) during training. Because the outcome of a lesion being present was relatively uncommon, future work should compare this interpretable baseline approach with class-weighted or resampled models and assess calibration in external cohorts. Although we prioritize care for patients from minoritized backgrounds, we chose to not explicitly encode race as a classifier variable to enhance robustness. We plan to carefully monitor the effect of the model on relevant subgroups throughout the deployment process. Furthermore, we hope that future versions will also make use of other data types, including biomarkers (e.g., serum neurofilament light chain^8^), to improve the accuracy of model predictions. We assessed the stability of our approach by adding an updated tranche of participant data (approximately a year of visits) to our initial 2017-2024 sample, which resulted in similar output performance of the model after retraining. Additional evaluations may be performed by replicating our approach in another clinical population such as MS PATHS^5^ and DISCO^9^.

Our initial algorithm needs further validation with larger data sets and prospective studies, which our tools and system infrastructure is ready to support. The operating threshold that was chosen was based on data from this study and could be different once it is studied prospectively. One component of our future approach is to consider both algorithmic and implementation bias as tool implementation is studied. Sociodemographic information is available in the EHR to support these analyses, including patient sex at birth and gender, race, insurance data, and geocode-based determinants of health. A limitation worth considering is that the current algorithm relies on data availability from sources that need to be manually entered by a clinician. If the MS Smartform is incomplete or incorrect, algorithm training is affected, and results may not be as meaningful or interpretable.

Additionally, our implementation design needs to consider integrating with the EHR and also how the interpretation of results are reported to the clinician. User-centered design concepts, such as transparency of information, need to be taken into account. For example, in addition to seeing the tool’s recommendation about the following scan they may want to access the patient-specific reasons (features) behind it from the model perspective. The availability of such detailed information would allow the clinician to assess if those aspects are consistent with the patient’s history and evidence, eventually building trust in the tool and the implied recommendations about whether or not to order brain imaging. Our algorithm could enable clinicians to adjust the length of time between MRIs, potentially reducing imaging frequency while improving individualized patient care. This algorithm could also be incorporated into care by non-MS specialists or doctors outside tertiary medical centers to better support individual patients. A key advantage of this approach is that it relies exclusively on routinely collected clinical and imaging variables available in most MS clinics, enabling eventual implementation without specialized imaging pipelines or advanced computational infrastructure.

In conclusion, our framework produced a pilot algorithm to help clinicians triage patients that would benefit from a subsequent surveillance MRI versus those who could be good candidates for decreased frequency of imaging. Once refined into a clinical decision-support tool, this approach may help support clinician-patient discussions regarding the value of image monitoring based on their individual patient disease characteristics.

## Data Availability

Data for this study can be requested from the corresponding authors upon reasonable request, consistent with the institutional review board and related guidelines and with appropriate data use agreements in place.

## Notes

### Competing Interest Statement

Dr. Sotirchos has received personal compensation for serving as a Consultant for Ad Scientiam. Dr. Sotirchos has received personal compensation for serving on a Scientific Advisory or Data Safety Monitoring board for Viela Bio. Dr. Sotirchos has received personal compensation for serving on a Scientific Advisory or Data Safety Monitoring board for Genentech. Dr. Sotirchos has received personal compensation for serving on a Scientific Advisory or Data Safety Monitoring board for Alexion. Dr. Sotirchos has received personal compensation for serving as an Ad Hoc Reviewer with National Institutes of Health. Dr. Mowry has received royalty payments from UpToDate. Dr. Mowry has received research support from Biogen and Genentech, editorial royalties from UpToDate, and consulting fees from BeCareLink, LLC. Dr. Peter Calabresi has received personal compensation as a consultant for Novartis. Dr. Peter Calabresi has received personal compensation for serving on a Scientific Advisory or Data Safety Monitoring board for Lilly. Dr. Calabresi has received personal compensation for serving on a Scientific Advisory or Data Safety Monitoring board for Idorsia. An immediate family member of Dr. Calabresi has received personal compensation for serving on a Scientific Advisory or Data Safety Monitoring board for MyMD. Dr. Calabresi has received personal compensation for serving as an Editor, Associate Editor, or Editorial Advisory Board Member and as a Grant reviewer for Myelin Repair Foundation. Dr. Calabresi has received publishing royalties from a publication relating to health care. Dr. Calabresi has received personal compensation for serving as a Study Section Member with NIH. Dr. Calabresi has received personal compensation for serving as a Speaker for CME with NYAS. Dr. Calabresi has received personal compensation for serving as a Speaker with Academic CME. Dr. Calabresi has received personal compensation for consulting with Lilly and Project Efflux. The institution of Dr. Calabresi has received research support from Genentech. The institution of Dr. Sotirchos has received research support from National Institutes of Health. The institution of Dr. Sotirchos has received research support from National Multiple Sclerosis Society. The institution of Dr. Sotirchos has received research support from Sumaira Foundation. The institution of Dr. Sotirchos has received research support from Genentech. Dr. Ferryman is a Member of the Digital Ethics Advisory Panel for Merck KGaA (Merck Germany) and a Member of the Institutional Review Board for the All of Us Research Program, National Institutes of Health.

### Funding Statement

This study was partially funded by JHU internal research investments as well as NIH K23NS117883 (ESS), NMSS RG-1904-33834 (ESS), NIH UM1NS132250 (WGR) and NMSS Sylvia Lawry Fellowship (KGR)

### Author Declarations

The IRB of Johns Hopkins Medicine gave ethical approval for this work.

### Summary of Updates

This has been revised to provide additional clarity around approaches, methods, and results.

